# Viral and host factors associated with SARS-CoV-2 disease severity in Georgia, USA

**DOI:** 10.1101/2023.10.25.23297530

**Authors:** Ludy R. Carmola, Allison Dorothy Roebling, Dara Khosravi, Rose M. Langsjoen, Andrei Bombin, Bri Bixler, Alex Reid, Cara Chen, Ethan Wang, Yang Lu, Ziduo Zheng, Rebecca Zhang, Phuong-Vi Nguyen, Robert A. Arthur, Eric Fitts, Dalia Arafat Gulick, Dustin Higginbotham, Azmain Taz, Alaa Ahmed, John Hunter Crumpler, Colleen Kraft, Wilbur A. Lam, Ahmed Babiker, Jesse J. Waggoner, Kyle P. Openo, Laura M. Johnson, Adrianna Westbrook, Anne Piantadosi

## Abstract

While SARS-CoV-2 vaccines have shown strong efficacy, their suboptimal uptake combined with the continued emergence of new viral variants raises concerns about the ongoing and future public health impact of COVID-19. We investigated viral and host factors, including vaccination status, that were associated with SARS-CoV-2 disease severity in a setting with low vaccination rates. We analyzed clinical and demographic data from 1,957 individuals in the state of Georgia, USA, coupled with viral genome sequencing from 1,185 samples. We found no difference in disease severity between individuals infected with Delta and Omicron variants among the participants in this study, after controlling for other factors, and we found no specific mutations associated with disease severity. Compared to those who were unvaccinated, vaccinated individuals experienced less severe SARS-CoV-2 disease, and the effect was similar for both variants. Vaccination within 270 days before infection was associated with decreased odds of moderate and severe outcomes, with the strongest association observed at 91-270 days post-vaccination. Older age and underlying health conditions, especially immunosuppression and renal disease, were associated with increased disease severity. Overall, this study provides insights into the impact of vaccination status, variants/mutations, and clinical factors on disease severity in SARS-CoV-2 infection when vaccination rates are low. Understanding these associations will help refine and reinforce messaging around the crucial importance of vaccination in mitigating the severity of SARS-CoV-2 disease.

## INTRODUCTION

Vaccinations against SARS-CoV-2 have undeniably demonstrated high efficacy in preventing COVID-19 infections and improving disease outcomes^1–3^, but their impact is challenged by the emergence of new variants carrying immune evasion mutations and the waning of immune responses over time^4^. Partly due to these issues, public trust in COVID-19 vaccines has diminished with each round of booster recommendations, especially in the southeast United States (US)^5^. Throughout the SARS-CoV-2 pandemic, the state of Georgia has consistently held one of the lowest rates of vaccine coverage in the nation^6^. One important approach to addressing vaccine hesitancy is to conduct studies – and disseminate their results – to populations with low vaccine uptake.

Understanding the impact of vaccination on SARS-CoV-2 disease outcomes requires consideration of other host factors such as underlying health conditions, sex, race, and socioeconomic factors, which impact SARS-CoV-2 disease severity ^7–17^. Viral factors are also important, especially as variants emerge with distinctive properties affecting pathogenesis and immune evasion. For example, the Delta variant has been associated with high risk of ICU admission and mortality^11,18,19^, while Omicron shows low neutralization sensitivity to vaccine induced immunity^20^.

In order to elucidate viral and host factors that contribute to disease outcomes in the context of low vaccination rates, we analyzed demographic and clinical data from 1,957 individuals in the state of Georgia. We also analyzed full viral genome sequences from 1,185 of these individuals to examine the influence of variants and mutations on disease severity. We leveraged a large study population and extensive demographic, clinical, and sequence data to define factors associated with SARS-CoV-2 disease severity in a region marked by low vaccination rates.

## RESULTS

### Clinical and demographic factors differ by SARS-CoV-2 vaccine status

Between May 2021 and May 2022, we identified 1,957 individuals who tested positive for SARS-CoV-2 within the Emory Healthcare system in Atlanta, Georgia. The majority of participants (66%) were residents of the Metro Atlanta area (Table 1, Figure 1A). The median age was 51 years (Interquartile range [IQR]=36,65), and 56% of individuals were female. The racial distribution of participants was predominantly Black (58%) or White (31%). Individuals experienced a range of clinical presentations and outcomes, from asymptomatic infection (15%) to death (2.8%) (Table 2). Among the 967 individuals in this study who were hospitalized, 625 (65%) of the hospitalizations were due to COVID-19.

**Figure 1.**
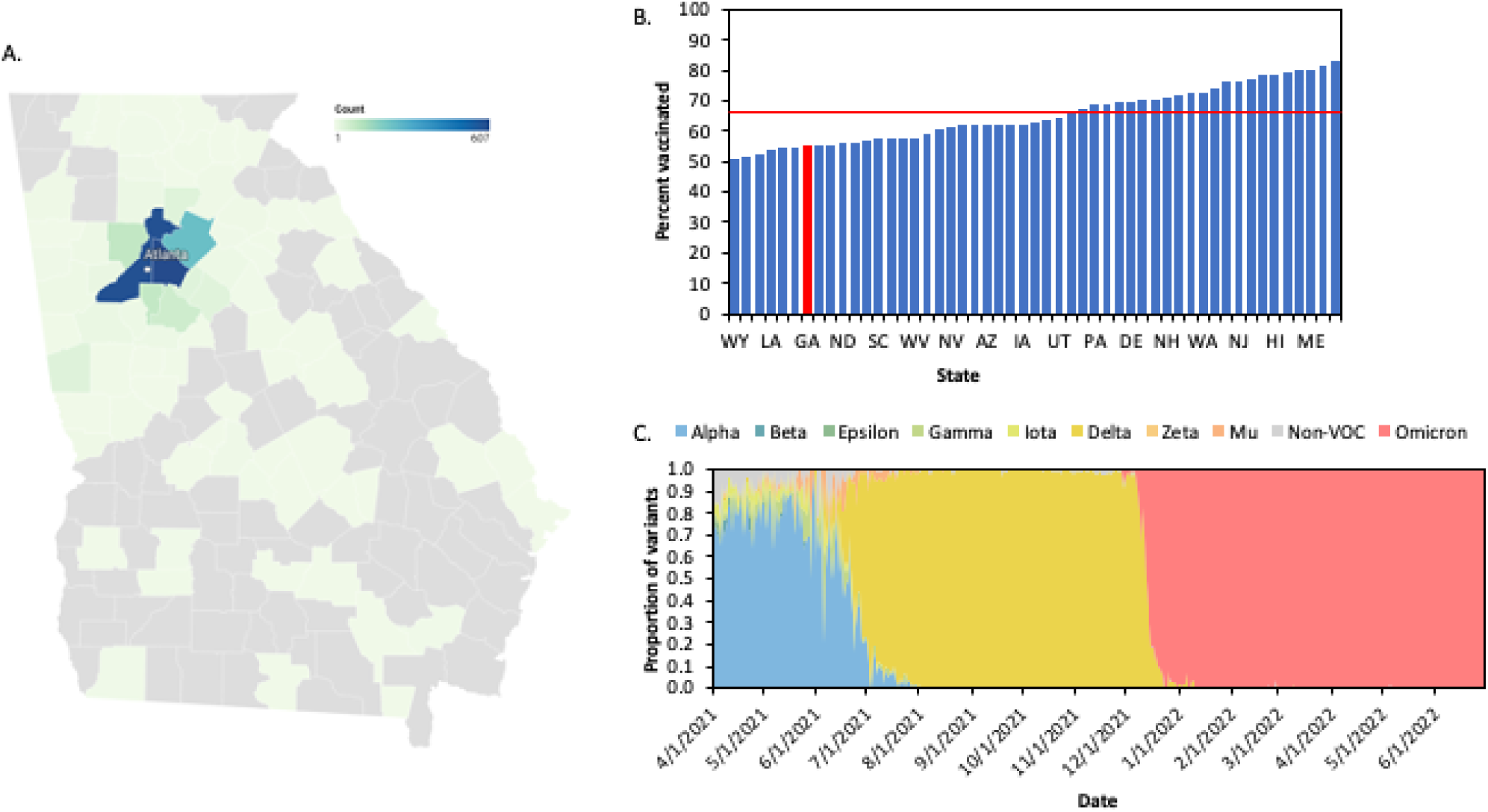
COVID-19 cases from May 2021-May 2022 in Georgia, US. **A**. 1,957 Emory Healthcare COVID-19 case mapped by counties in and around Atlanta, GA. **B**. Percent of population fully vaccinated against SARS-CoV-2 by May 2022 in each state in the United States. Red line represents national average (66.7%). Red bar represents Georgia. **C.** Proportion of variants circulating in Georgia from April 2021-June 2022. Sequences obtained from GISAID.

**Table 1.**
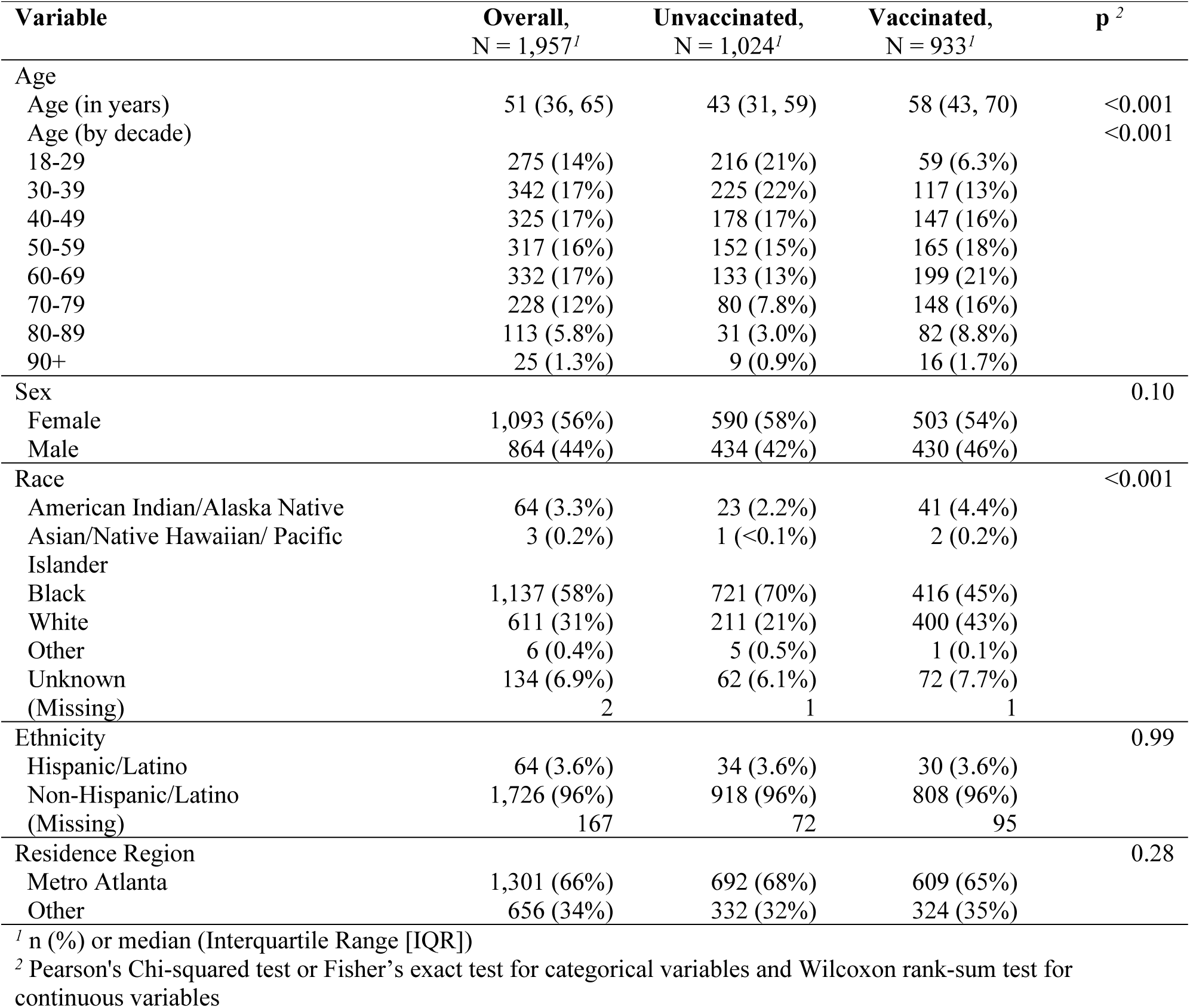
Demographic Characteristics by Vaccination Status.

**Table 2.**
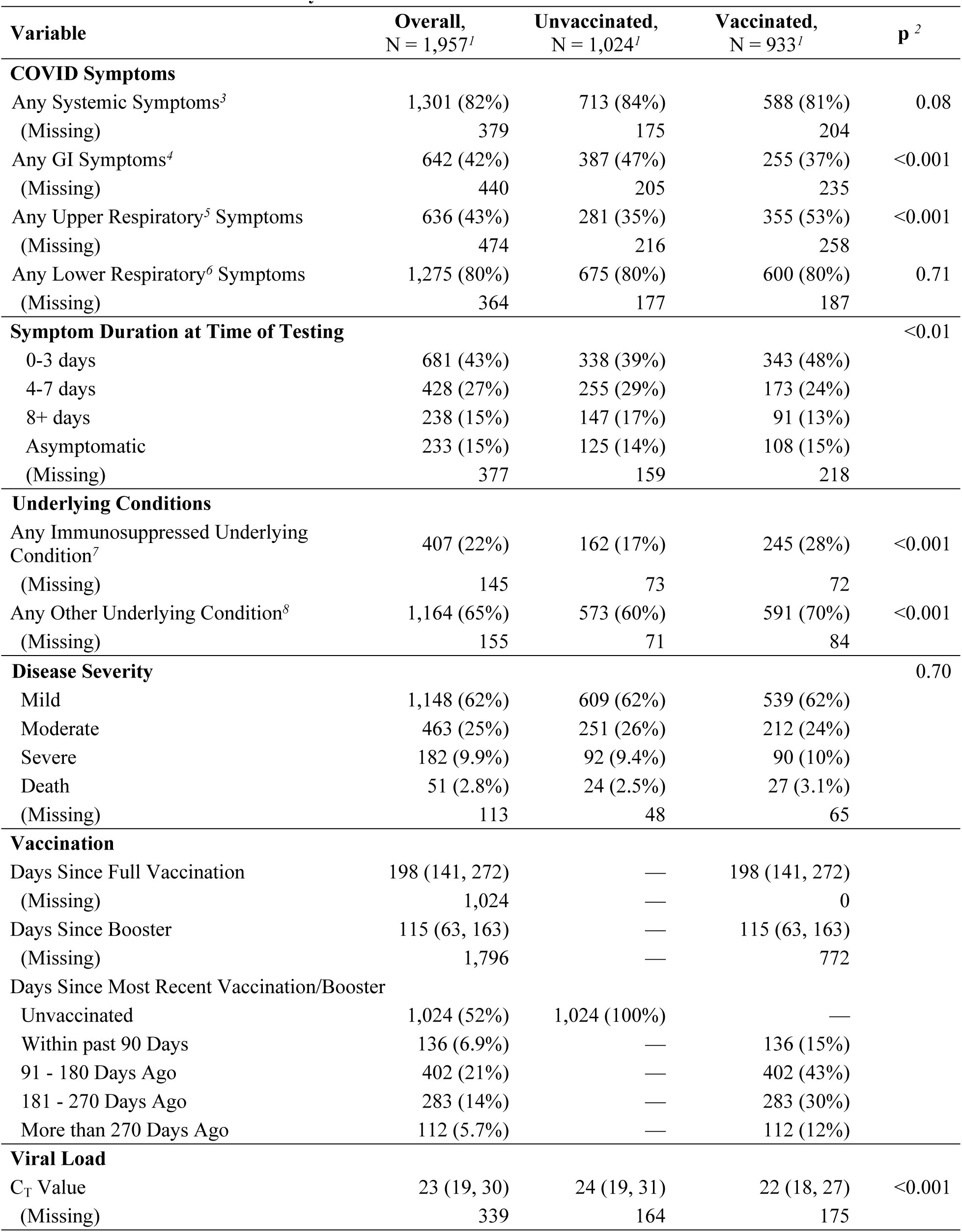
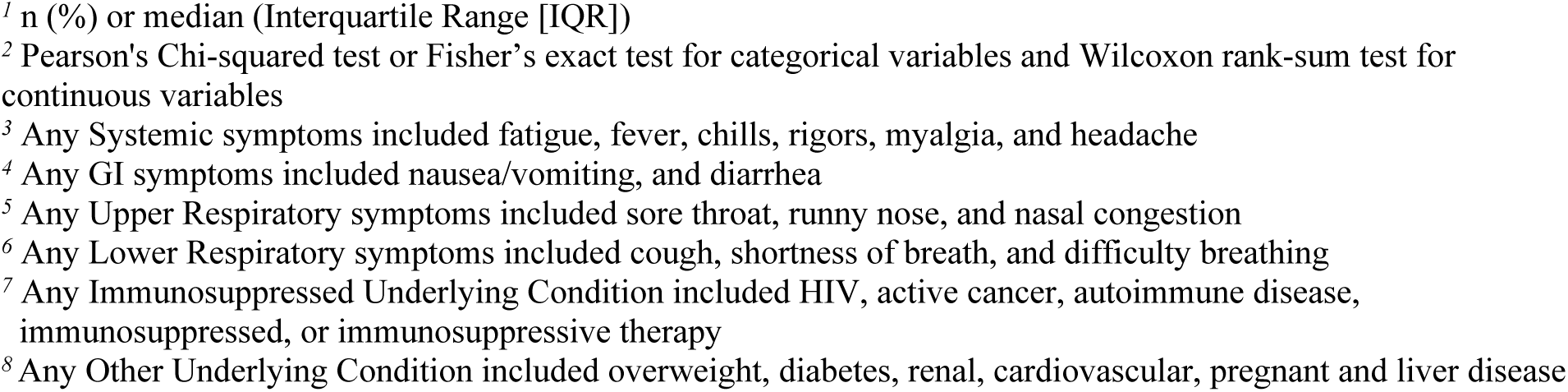
Clinical Characteristics by Vaccination Status.

| **Variable** | **Overall,**<br>**N = 1,957<sup>i</sup>** | **Unvaccinated,**<br>**N = 1,024<sup>i</sup>** | **Vaccinated,**<br>**N = 933<sup>i</sup>** | **p<sup>2</sup>** |
| --- | --- | --- | --- | --- |
| **COVID Symptoms** |  |  |  |  |
| Any Systemic Symptoms<sup>3</sup> | 1,301 (82%) | 713 (84%) | 588 (81%) | 0.08 |
| (Missing) | 379 | 175 | 204 |  |
| Any GI Symptoms<sup>4</sup> | 642 (42%) | 387 (47%) | 255 (37%) | <0.001 |
| (Missing) | 440 | 205 | 235 |  |
| Any Upper Respiratory<sup>5</sup> Symptoms | 636 (43%) | 281 (35%) | 355 (53%) | <0.001 |
| (Missing) | 474 | 216 | 258 |  |
| Any Lower Respiratory<sup>6</sup> Symptoms | 1,275 (80%) | 675 (80%) | 600 (80%) | 0.71 |
| (Missing) | 364 | 177 | 187 |  |
| **Symptom Duration at Time of Testing** |  |  |  | <0.01 |
| 0-3 days | 681 (43%) | 338 (39%) | 343 (48%) |  |
| 4-7 days | 428 (27%) | 255 (29%) | 173 (24%) |  |
| 8+ days | 238 (15%) | 147 (17%) | 91 (13%) |  |
| Asymptomatic | 233 (15%) | 125 (14%) | 108 (15%) |  |
| (Missing) | 377 | 159 | 218 |  |
| **Underlying Conditions** |  |  |  |  |
| Any Immunosuppressed Underlying Condition<sup>7</sup> | 407 (22%) | 162 (17%) | 245 (28%) | <0.001 |
| (Missing) | 145 | 73 | 72 |  |
| Any Other Underlying Condition<sup>8</sup> | 1,164 (65%) | 573 (60%) | 591 (70%) | <0.001 |
| (Missing) | 155 | 71 | 84 |  |
| **Disease Severity** |  |  |  | 0.70 |
| Mild | 1,148 (62%) | 609 (62%) | 539 (62%) |  |
| Moderate | 463 (25%) | 251 (26%) | 212 (24%) |  |
| Severe | 182 (9.9%) | 92 (9.4%) | 90 (10%) |  |
| Death | 51 (2.8%) | 24 (2.5%) | 27 (3.1%) |  |
| (Missing) | 113 | 48 | 65 |  |
| **Vaccination** |  |  |  |  |
| Days Since Full Vaccination | 198 (141, 272) | — | 198 (141, 272) |  |
| (Missing) | 1,024 | — | 0 |  |
| Days Since Booster | 115 (63, 163) | — | 115 (63, 163) |  |
| (Missing) | 1,796 | — | 772 |  |
| Days Since Most Recent Vaccination/Booster |  |  |  |  |
| Unvaccinated | 1,024 (52%) | 1,024 (100%) | — |  |
| Within past 90 Days | 136 (6.9%) | — | 136 (15%) |  |
| 91 - 180 Days Ago | 402 (21%) | — | 402 (43%) |  |
| 181 - 270 Days Ago | 283 (14%) | — | 283 (30%) |  |
| More than 270 Days Ago | 112 (5.7%) | — | 112 (12%) |  |
| **Viral Load** |  |  |  |  |
| C<sub>T</sub> Value | 23 (19, 30) | 24 (19, 31) | 22 (18, 27) | <0.001 |
| (Missing) | 339 | 164 | 175 |  |

<sup>1</sup> n (%) or median (Interquartile Range [IQR])
<sup>2</sup> Pearson's Chi-squared test or Fisher's exact test for categorical variables and Wilcoxon rank-sum test for continuous variables
<sup>3</sup> Any Systemic symptoms included fatigue, fever, chills, rigors, myalgia, and headache
<sup>4</sup> Any GI symptoms included nausea/vomiting, and diarrhea
<sup>5</sup> Any Upper Respiratory symptoms included sore throat, runny nose, and nasal congestion
<sup>6</sup> Any Lower Respiratory symptoms included cough, shortness of breath, and difficulty breathing
<sup>7</sup> Any Immunosuppressed Underlying Condition included HIV, active cancer, autoimmune disease, immunosuppressed, or immunosuppressive therapy
<sup>8</sup> Any Other Underlying Condition included overweight, diabetes, renal, cardiovascular, pregnant and liver disease

During the period of this study, the state of Georgia had the 7^th^ lowest vaccination rate in all 50 United States and the District of Columbia, with 55.1% of the population vaccinated (Figure 1B). In our study, a slightly lower proportion (48%) of individuals were vaccinated, in part based our study was designed to ensure inclusion of unvaccinated individuals. Vaccinated individuals were significantly older than unvaccinated individuals (median age of 58 years vs 43 years, p<0.001) (Table 1). We observed a large disparity in vaccine status by race; among the unvaccinated participants, 70% were Black and 21% White, while among the vaccinated participants, 45% were Black and 43% were White (Table 1). Vaccination status was not significantly associated with any other demographic variable evaluated.

We found that vaccinated individuals were more likely to have underlying medical comorbidities than unvaccinated individuals. Notably, 28% of vaccinated individuals were immunocompromised, compared to 17% of unvaccinated individuals (p <0.001, Table 2). Vaccinated individuals were also more likely to have hypertension (56% vs. 39%, p <0.001), cardiovascular disease (36% vs. 23%, p <0.001), diabetes (28% vs. 19%, p <0.001), renal disease (24% vs. 11%, p <0.001), and autoimmune disease (7% vs. 4%, p = 0.03) (Table S1). Vaccinated individuals were less likely to be pregnant (1% vs. 6%, p <0.001) (Table 2, Table S1), however, at the time of the study, vaccines were not yet approved for pregnant individuals. These findings are consistent with higher rates of vaccination in individuals with medical comorbidities.

We also found differences in clinical symptoms by vaccination status. Compared to unvaccinated individuals, those who were vaccinated were less likely to have fever (52% vs. 42%, p <0.001), chills (38% vs. 32%, p = 0.02), nausea/vomiting (35% vs. 25%, p <0.001), and shortness of breath or difficulty breathing (49% vs. 39%, p <0.001); they were more likely to have sore throat (17% vs. 22%, p = 0.02) and runny nose/nasal congestion (27% vs. 45%, p <0.001) (Table S1). These findings are consistent with milder disease in vaccinated individuals.

SARS-CoV-2 C_T_ value by qRT-PCR is often used as a rough proxy for viral load, and we found several key factors associated with SARS-CoV-2 C_T_. Because multiple qRT-PCR assays were used within the Emory Healthcare system during this time, we controlled for assay variability (Table 3). Vaccinated individuals had a slightly lower C_T_ than those who were unvaccinated (–0.70, SE=0.26, p =0.01). Though this finding seems counterintuitive, we attribute it to vaccinated individuals presenting for testing earlier after symptom onset, compared to unvaccinated individuals (**Table 2**). Symptom duration was significantly inversely associated with C_T_ (Table 3). Specifically, after controlling for other variables, and compared to asymptomatic individuals, those who were tested within 0-3 days after symptom onset had a lower C_T_ value by 3.0 cycles (Standard Error [SE]=0.55, p <0.001) and those who were tested within 4-7 days had a lower C_T_ value by 1.62 cycles (SE=0.61, p = 0.01). Those who were tested 8 or more days from symptom onset did not have a significantly different C_T_ than those who were asymptomatic (p = 0.77) (Table 3). No significant differences in C_T_ were observed between variants, after adjusting for other factors (Table 3).

**Table 3.** Association between C_T_ value and clinical factors.

| **Variable** | **Beta** | **SE** | **p** |
| --- | --- | --- | --- |
| Vaccination status |  |  |  |
| Unvaccinated | Ref | — | — |
| Vaccinated | -0.70 | 0.26 | 0.01 |
| Lineage |  |  |  |
| Alpha | Ref | — | — |
| Delta | -0.23 | 0.87 | 0.79 |
| Omicron | 1.68 | 0.92 | 0.07 |
| Symptom Duration at Time of Testing |  |  |  |
| Asymptomatic | Ref | — | — |
| 0-3 days | -2.99 | 0.55 | <0.001 |
| 4-7 days | -1.62 | 0.61 | 0.01 |
| 8+ days | 0.20 | 0.68 | 0.77 |
| Assay |  |  |  |
| Cepheid Gene Xpert | Ref | — | — |
| BioFire Defense, LLC - BioFire COVID-19 Test | -1.78 | 2.82 | 0.54 |
| Cepheid - Xpert Xpress SARS-CoV-2/Flu/RSV | -0.39 | 2.85 | 0.89 |
| Roche Cobas | -0.56 | 0.35 | 0.11 |
| Other | 4.37 | 7.81 | 0.59 |

SE = Standard Error, Ref = Reference Level

### Variant frequency differs between vaccinated and unvaccinated individuals

We sequenced full SARS-CoV-2 genomes from residual nasopharyngeal swab samples from 1,185 individuals. The minimum genome coverage of the samples was 76% and the median sequencing depth was 1,804 (Supplementary Data File). Sequences were primarily Delta (68%) and the BA.1 sublineage of the Omicron variant (23%), followed by other Omicron sublineages (5.1%), Alpha (2.5%) and less than 1% each of Beta, Gamma, Mu, A.2.5, and B.1 (Table 4). In Georgia, during the time of this study (May 2021-May 2022), Delta accounted for 55% of infections, Omicron for 40%, and Alpha for 2.5% (Figure 1C, Table S2). Therefore, our study included a somewhat higher proportion of Delta and a lower proportion of Omicron than was circulating in the state. The distribution of all other variants aligned with the overall variant distribution observed in Georgia (Table S2, Table 4).

**Table 4.** SARS-CoV-2 Variants by Vaccination Status.

| **Variable** | **Overall,**<br>**N = 1,957<sup>1</sup>** | **Unvaccinated,**<br>**N = 1,024<sup>1</sup>** | **Vaccinated,**<br>**N = 933<sup>1</sup>** | **p<sup>2</sup>** |
| --- | --- | --- | --- | --- |
| Variant |  |  |  | <0.001 |
| A.2.5 | 1 (<0.1%) | 0 (0%) | 1 (0.2%) |  |
| Alpha | 30 (2.5%) | 24 (4.0%) | 6 (1.0%) |  |
| B.1 | 1 (<0.1%) | 0 (0%) | 1 (0.2%) |  |
| Beta | 1 (<0.1%) | 0 (0%) | 1 (0.2%) |  |
| Delta | 805 (68%) | 415 (70%) | 390 (66%) |  |
| Gamma | 6 (0.5%) | 5 (0.8%) | 1 (0.2%) |  |
| Mu | 4 (0.3%) | 4 (0.7%) | 0 (0%) |  |
| Omicron | 337 (28%) | 146 (25%) | 191 (32%) |  |
| (Missing) | 772 | 430 | 342 |  |

The distribution of SARS-CoV-2 variants in our study was different for vaccinated and unvaccinated individuals (p <0.001) (Table 4). Omicron had a higher frequency in vaccinated individuals (32%) than unvaccinated (25%), whereas Alpha and Delta were more common in unvaccinated compared to vaccinated individuals (Table 4). These differences correspond with the timing of each variant’s circulation compared to vaccine rollout, though decreased vaccine effectiveness against Omicron may also contribute^21^.

In addition to the frequency of VOCs among vaccinated and unvaccinated individuals, we investigated the frequency of individual mutations. Within each VOC – Alpha, Delta, and Omicron – no sequence characteristics, including mutations, deletions, and insertions, were different between viruses infecting vaccinated and unvaccinated individuals (Figure 2, Figure S1). However, the number of non-lineage defining mutations was lower in vaccinated compared to unvaccinated individuals (Figure 1D) suggesting that vaccination may have an impact on viral diversity within a host. Phylogenetic analysis demonstrated that sequences from vaccinated individuals were intermixed with sequences from unvaccinated individuals, further confirming no distinct features of post vaccination infections (Figure S2).

**Figure 2.**
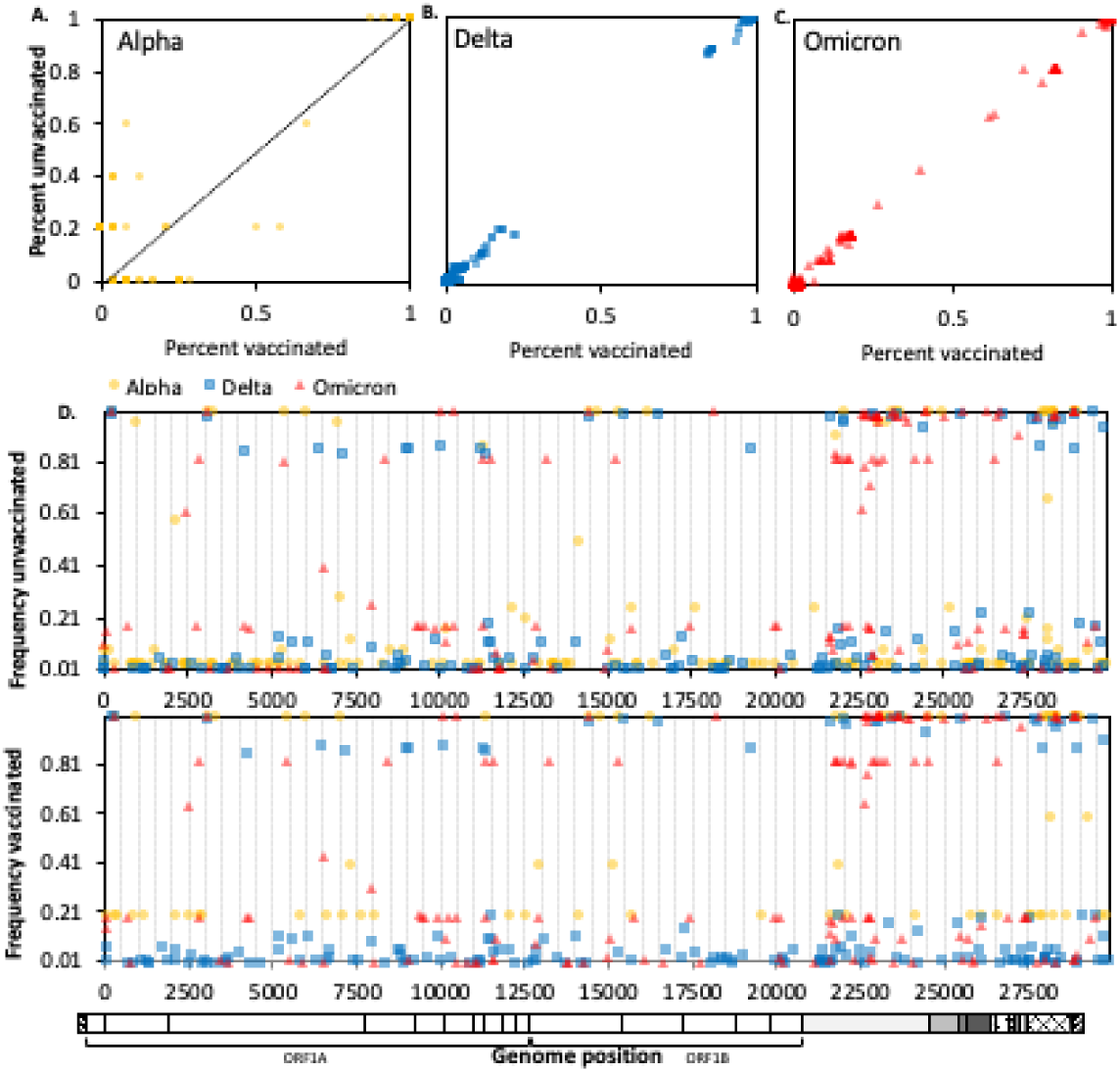
Frequencies of single nucleotide polymorphisms (SNPs) among SARS-COV-2 genome sequences from vaccinated and unvaccinated individuals. Each point represents a single SNP plotted by its frequency in sequences from unvaccinated individuals (Y-axis) versus its frequency in sequences from vaccinated individuals (X-axis). Data is divided by WHO variant classifications Alpha **(A)**, Delta **(B)**, and Omicron **(C)**. Mutations observed along the diagonal depict mutations observed equally among vaccinated and unvaccinated individuals. Mutations observed moving away from the diagonal represent mutations observed in either vaccinated (X-axis) or unvaccinated (Y-axis) individuals. (**D**) Frequency of SNPs in SARS-CoV-2 sequences from unvaccinated (top) and vaccinated (bottom) individuals, by genome position (x-axis). In all panels, Alpha is represented by yellow circles, Delta by blue squares, and Omicron by red triangles.

In-depth metagenomic analysis of 513 samples did not reveal any viral co-infections (Supplementary Data File).

### Age, underlying conditions, and vaccination status are associated with disease severity

We evaluated associations between disease severity and demographic characteristics, underlying health conditions, vaccination status, and SARS-CoV-2 variant. Disease severity was defined according to the WHO clinical progression scale^22^. Mild disease included asymptomatic infection and symptomatic infection without hospitalization. Moderate disease included hospitalized individuals without oxygen therapy or oxygen by mask or nasal prongs. Severe disease included the use of oxygen by noninvasive or high flow, intubation and mechanical ventilation, vasopressors, dialysis, or extracorporeal membrane oxygenation. Death included in-hospital deaths directly linked to COVID.

Using an adjusted multinomial logistic regression model that included demographic characteristics, underlying conditions, vaccination status and variant, we found that age, certain underlying medical conditions, and time since most recent vaccination were significantly associated with disease severity (Table 5). Underlying health conditions significantly associated with disease severity included chronic lung disease, renal disease, and the use of systemic immunosuppressive therapy prior to hospitalization. Other conditions showed weak or non-significant associations with disease severity, including pregnancy, diabetes, liver disease, and autoimmune disease. The Omicron variant did not show a significant association with disease severity compared to the Delta variant in the adjusted model although an association was observed in an unadjusted model (Table 5, Table S3). Finally, compared to unvaccinated individuals, vaccination within 91-180 days (about 3 – 6 months) was associated with lower odds of moderate disease, severe disease, and death (Table 5). There were similar effects across all disease severity outcomes when vaccination occurred between 181 – 270 days (about 6 to 9 months). When vaccination occurred more than 270 days ago, however, there were no significant differences in disease severity when compared to the unvaccinated group. Additionally, vaccination that had occurred within the past 90 days (about 3 months) was associated with lower odds of moderate disease relative to mild disease but was not associated with lower odds of severe disease or death. In short, vaccinations were most protective against moderate disease, severe disease, and death when they occurred within the prior 3-9 months. Overall, after controlling for multiple host and viral factors, we found that age, chronic lung disease, renal disease, and immunosuppressive therapy increased the odds of progressively more severe COVID-19 disease, while vaccination decreased the odds and SARS-CoV-2 variant had no effect.

**Table 5.** Adjusted Multinomial Logistic Regression Models Testing the Association between Demographics, Underlying Health Conditions, and COVID-Related Characteristics with Disease.

| Variable | Moderate<sup>1</sup> |  |  | Severe<sup>1</sup> |  |  | Death<sup>1</sup> |  |  |
| --- | --- | --- | --- | --- | --- | --- | --- | --- | --- |
|  | aOR<sup>2</sup> | 95% CI<sup>2</sup> | p | aOR<sup>2</sup> | 95% CI<sup>2</sup> | p | aOR<sup>2</sup> | 95% CI<sup>2</sup> | p |
| *Demographics* |  |  |  |  |  |  |  |  |  |
| Age | **1.05** | **1.03, 1.06** | **<0.001** | **1.05** | **1.03, 1.07** | **<0.001** | **1.09** | **1.05, 1.13** | **<0.001** |
| Male | 1.28 | 0.89, 1.85 | 0.18 | 1.34 | 0.82, 2.19 | 0.24 | 1.21 | 0.52, 2.81 | 0.66 |
| White | 0.70 | 0.46, 1.06 | 0.09 | 0.89 | 0.51, 1.54 | 0.67 | 0.73 | 0.27, 1.96 | 0.53 |
| *Underlying Conditions* |  |  |  |  |  |  |  |  |  |
| Chronic Lung Disease | **2.10** | **1.38, 3.17** | **<0.001** | **1.98** | **1.15, 3.41** | **0.01** | 1.31 | 0.50, 3.46 | 0.58 |
| Hypertension | 1.02 | 0.65, 1.59 | 0.94 | 0.97 | 0.53, 1.76 | 0.91 | 3.46 | 0.88, 13.57 | 0.08 |
| Overweight | 1.40 | 0.95, 2.07 | 0.09 | 1.17 | 0.69, 1.99 | 0.56 | 1.61 | 0.65, 4.01 | 0.30 |
| Cardiovascular Disease | **1.63** | **1.06, 2.50** | **0.03** | 1.45 | 0.82, 2.55 | 0.20 | 1.08 | 0.44, 2.63 | 0.87 |
| Diabetes | 0.99 | 0.64, 1.54 | 0.98 | 1.45 | 0.83, 2.54 | 0.20 | 1.09 | 0.43, 2.74 | 0.86 |
| Renal Disease | **2.64** | **1.61, 4.32** | **<0.001** | **1.91** | **1.01, 3.64** | **0.05** | **3.63** | **1.41, 9.36** | **0.01** |
| Liver Disease | 1.13 | 0.48, 2.69 | 0.78 | 1.84 | 0.69, 4.92 | 0.22 | 1.48 | 0.25, 8.78 | 0.67 |
| Autoimmune Disease | 1.78 | 0.87, 3.65 | 0.11 | 1.95 | 0.77, 4.95 | 0.16 | 3.96 | 1.00, 15.65 | 0.05 |
| Immunocompromised<sup>3</sup> | 1.08 | 0.55, 2.11 | 0.82 | 0.59 | 0.25, 1.40 | 0.23 | 3.26 | 0.83, 12.71 | 0.09 |
| Systemic Immunosuppressive Therapy or Meds. | **3.45** | **1.88, 6.32** | **<0.001** | **5.01** | **2.38, 10.51** | **<0.001** | 1.48 | 0.38, 5.71 | 0.57 |
| *Days Since Most Recent Vaccination/Booster* |  |  |  |  |  |  |  |  |  |
| Unvaccinated | Ref | — |  | Ref | — |  | Ref | — |  |
| Within past 90 Days<sup>4</sup> | **0.37** | **0.16, 0.88** | **0.02** | 0.40 | 0.13, 1.26 | 0.12 | 0.72 | 0.18, 2.86 | 0.64 |
| 91 - 180 Days Ago<sup>4</sup> | **0.32** | **0.19, 0.54** | **<0.001** | **0.42** | **0.21, 0.82** | **0.01** | **0.31** | **0.10, 0.96** | **0.04** |
| 181 - 270 Days Ago<sup>4</sup> | **0.32** | **0.18, 0.56** | **<0.001** | **0.38** | **0.17, 0.80** | **0.01** | **0.14** | **0.04, 0.59** | **0.01** |
| More than 270 Days Ago | 0.53 | 0.23, 1.21 | 0.13 | 0.88 | 0.34, 2.29 | 0.79 | 0.34 | 0.06, 1.95 | 0.23 |
| *Lineage* |  |  |  |  |  |  |  |  |  |
| Delta | Ref | — |  | Ref | — |  | Ref | — |  |
| Omicron<sup>5</sup> | 0.95 | 0.61, 1.47 | 0.81 | 1.42 | 0.81, 2.50 | 0.22 | 2.07 | 0.84, 5.12 | 0.12 |

<sup>1</sup> "Mild" is the Reference Category
<sup>2</sup> aOR = Adjusted Odds Ratio, CI = Confidence Interval, Ref = Reference Level
<sup>3</sup> HIV infection, active cancer, solid organ transplant, hematopoietic stem cell transplant
<sup>4</sup> Compared to unvaccinated individuals
<sup>5</sup> Compared to Delta

## DISCUSSION

Our comprehensive analysis of host and viral factors associated with SARS-CoV-2 disease severity in a setting with low vaccination rates led to several key findings. First, age and underlying health conditions – especially chronic lung disease, renal disease, and the use of immunosuppressive therapy – were associated with more severe disease and death. Second, SARS-CoV-2 variant and viral mutations were not associated with disease severity in this study population, which was comprised of individuals who sought medical care. And third, vaccination was protective against severe outcomes for both Delta and Omicron variants to a similar degree. Unique features of our study included the analysis of a large number of SARS-CoV-2 full viral genome sequences linked to extensive clinical and demographic data, and our focus on a relatively under-studied region of the U.S.

Georgia is an important proxy for the southeastern U.S. and other populations with high numbers of vaccine refusals, inequitable access to healthcare, and low insurance coverage^23–25^. Emphasizing the positive impact of SARS-CoV-2 vaccination among this population, similar to others in the U.S.^26,27^, is critical as new variants emerge. It is also important to note that among the individuals in this study who contracted SARS-CoV-2 after being vaccinated, a greater proportion reported milder upper respiratory symptoms like sore throat and runny nose, while a lower percentage experienced more severe symptoms such as nausea/vomiting, fever, and shortness of breath/difficulty breathing, in comparison to unvaccinated individuals. Vaccinations were most protective against severe COVID outcomes when they occurred within the prior 3-9 months. This finding has timely implications on a national level, given persistently low vaccine uptake, especially of the bivalent SARS-CoV-2 vaccine^5^, and the need for ongoing vaccine updates targeting emerging variants^4^. Results from our study will help emphasize the benefits of vaccination to the public as a means of safeguarding against severe COVID outcomes.

Our results also indicated the importance of demographic and clinical factors associated with SARS-CoV-2 disease severity, despite vaccination. Age was a key risk factor; after accounting for vaccination status, demographic factors, health conditions, and SARS-CoV-2 variant, our analysis revealed that for each additional year of age, the odds of experiencing more severe outcomes compared to mild disease increased by 5%. The association between age and disease severity has been consistently observed, particularly among individuals aged 65 and above^7–10^. Thus, relying solely on vaccination may be insufficient for reducing disease severity and mortality among older individuals. U.S. Census Bureau data indicates that the population is aging, with Georgians aging at an even faster rate^28^, underscoring the need for additional preventative and treatment measures.

In addition to age, we found that chronic lung disease, renal disease, and the use of immunosuppressive therapy also increased the odds of experiencing moderate and/or severe infection and/or death. In previous studies, cardiovascular disease, diabetes, chronic respiratory conditions, obesity, and compromised immune systems have also been found to increase the risk of severe illness^10–17^. Interestingly, we found that cardiovascular disease was only associated with moderate disease (without any association with severe disease or mortality), while diabetes and being overweight were not associated with disease severity in our final multivariate model. One explanation for this discrepancy could be that these two conditions serve as indicators for factors that we controlled in our study. It is noteworthy that when we did not adjust for any factors, these conditions were significantly associated with increased odds of severe disease.

It is surprising that we found no difference in disease severity between Delta and Omicron variants, since multiple prior studies have found that the risk of hospitalization, ICU admission, and mortality vary by variant^11,18,19,29^. This discrepancy may be due to the fact that all individuals in our study sought medical care, so we did not include individuals with minimal symptoms. Our observation that vaccination was similarly protective for individuals with Delta and Omicron is consistent with results from another recent study among hospitalized patients^30^.

We did not find viral factors associated with post-vaccine infection. Most post-vaccine infections were caused by the predominant lineage of the time. Within each variant (Delta and Omicron), no SARS-CoV-2 SNPs, deletions, or insertions were more common in vaccinated individuals than unvaccinated individuals. These results are different from a prior study of similar size, which found more resistance mutations (e.g. L452* and E484*) in vaccinated compared to unvaccinated individuals in the pre-Omicron era^16^. Our negative finding likely reflects the challenge of identifying the effect of an individual virus mutation in an increasingly complex immune landscape. Interestingly, we found that vaccinated individuals had fewer non-lineage-defining SNPs than unvaccinated individuals, suggesting less diversity and potentially less viral evolution within vaccinated individuals. This is consistent with a recent study investigating within-host genetic diversity of SARS-CoV-2 in unvaccinated and vaccinated individuals.^31^

Our study had several limitations: We only included individuals who presented to care, thus skewing our study population towards individuals with more severe disease than the general population. In addition, due to our study design, we could not collect reliable data regarding prior SARS-CoV-2 infection(s) thus, we were unable to account for natural immunity. Given the retrospective study design, there may be residual confounding, however we adjusted for important variables such age, demographics, pre-existing health conditions and vaccination status.

In summary, our findings underscore the critical role of vaccination status, age, and medical comorbidities – especially immunosuppression, chronic kidney disease, and chronic lung disease – in determining disease severity and outcomes among SARS-CoV-2 infected individuals, regardless of virus variant. We contribute valuable insights into the nuanced relationship between these factors, highlighting the importance of considering demographic, clinical, and genetic variables when evaluating disease severity. Ultimately, our results will be valuable in strengthening and reinforcing messaging around SARS-CoV-2 vaccination, especially in settings of low vaccine uptake.

## METHODS

### Clinical and demographic data

This study was approved by the institutional review board at Emory University under protocol STUDY00000260, with a waiver of consent. All positive SARS-CoV-2 samples from Emory University Hospital Molecular and Microbiology Laboratories collected between 5/3/21 and 5/31/22 were reviewed for inclusion in this study. In addition to symptomatic testing, SARS-CoV-2 tests were administered before admission or outpatient procedure as part of Emory Healthcare System’s universal SARS-CoV-2 screening. Individuals were considered vaccinated if they had received a complete vaccine series (2 doses of the Pfizer-BioNTech or Moderna vaccines or 1 dose of the Janssen vaccine) at least 14 days before their first positive test result. Individuals were excluded from the study if they were partially vaccinated or reported out-of-state residency. From May 2021-September 2021, individuals were included in the study on a case-match basis; for each post-vaccine case identified, 2-3 non-vaccinated control cases were selected at random from the positive samples tested in the same calendar week. From October 2021-May 2022, all SARS-CoV-2 positive individuals identified who met inclusion criteria were included.

The Centers for Disease Control and Prevention (CDC)-funded Georgia Emerging Infections Program (GA EIP) performs active, population– and laboratory-based surveillance for hospitalized cases of SARS-CoV-2 in metropolitan Atlanta, GA (population ∼4 million). Patient vaccination status was retrieved by GA EIP from the Georgia Registry of Immunization Transactions and Services (GRITS) database. State of residency was retrieved from Georgia’s State Electronic Notifiable Disease Surveillance System (SENDSS). Patient demographics, SARS-CoV-2 RT-PCR results, and C_T_ value, underlying medical conditions, symptomatic illness, hospitalization, and disease outcome were obtained from the electronic medical record (EMR). In constructing symptom categories, systemic symptoms were defined as fatigue, fever, chills, rigors, myalgia, and headache. Gastrointestinal symptoms were defined as nausea/vomiting and diarrhea. Upper respiratory symptoms were defined as sore throat, runny nose, and nasal congestion. Lower respiratory symptoms were defined as cough, and shortness of breath/difficulty breathing. Immunosuppressed was defined as HIV, active cancer, autoimmune disease, or immunosuppressive therapy. “Other” underlying conditions were defined as overweight, diabetes, renal, cardiovascular, pregnant, and liver disease.

Disease severity was defined according to the WHO clinical progression scale^22^. The scale includes mild disease-asymptomatic and symptomatic SARS-CoV-2 infection without hospitalization, moderate disease-hospitalization without oxygen therapy or hospitalization with oxygen by mask or nasal prongs, severe disease-hospitalization with use of oxygen by noninvasive or high flow, intubation and mechanical ventilation, vasopressors, dialysis, or extracorporeal membrane oxygenation, and death.

Data management and cleaning were conducted in Excel v16.73 and SAS studio v3.81.

### SARS-CoV-2 sequencing and analysis

Residual nasopharyngeal (NP) swab samples were obtained from the Emory University Hospital Molecular and Microbiology Laboratories. NP samples underwent RNA extraction, DNase treatment, and cDNA synthesis followed by metagenomic or amplicon-based library construction. For metagenomic sequencing, Nextera XT (Illumina) and Illumina sequencing were performed as previously described^32^. Amplicon-based sequencing was performed using the xGEN SARS-CoV-2 kit (IDT) as previously described^33^.

Reference-based SARS-CoV-2 genome assembly was performed using viral-ngs v2.1.12.0^34^ or Viralrecon^35^ for metagenomic and amplicon sequencing, respectively, with reference strain NC_045512. SARS-CoV-2 lineages were determined using Pangolin^36^. Sequences were aligned and visualized in Geneious Prime (https://www.geneious.com). Consensus-level single nucleotide polymorphisms (SNPs) and insertions/deletions were identified using the Nextstrain web-based mutation calling tool^37^.

For phylogenetic analysis, 411,634 reference sequences, collected between May 1, 2021, and May 31, 2022, were downloaded from NCBI and were aligned with our study sequences to reference strains Wuhan/Hu-1/2019 and Wuhan/WHO/2019 using Nextalign within the Nextstrain v3.2.4 pipeline 7. This dataset was subsampled in Nextstrain using a custom scheme, in which crowd penalty was set to 0.0 to select 1000 sequences most genetically similar to our sequence dataset. Maximum likelihood phylogenetic trees were constructed using default settings of the Nextstrain SARS-CoV-2 Workflow with TreeTime v0.8.6 ^38^.

### Viral metagenomic analysis

To assess the presence of viral co-infections in 513 samples that underwent metagenomic sequencing, reads were first passed through a pre-processing pipeline including deduplication with Clumpify.sh in the BBMap tools (https://sourceforge.net/projects/bbmap/). Deduplicated reads were trimmed with Trimmomatic Version 0.40 and filtered for quality, with flags leading:3, trailing:3, slidingwindow:4:15, minlen:36 (https://github.com/usadellab/Trimmomatic). Pre-processed reads were run through kraken2 v2.1.3 against the k2_pluspf_20210127 database to assign each read to a taxonomic group, then adjusted for significance with Bracken. Within the Kraken Tools packages, the extract_kraken_reads.py script was used to separate reads by taxonomic ID for human taxID_hg=“9606”, bacteria taxID_bac=“2”, fungus taxID_fungus=“4751”, viruses taxID_virus=“10239”, and COVID-19 taxID_COVID=“2697049”. Custom shell and R scripts were used to determine if the following viruses were found in each sample: Human mastadenovirus C taxID=129951, Coronavirus HKU1 taxID=443239, Coronavirus NL63 taxID=277944, Coronavirus 299E taxID=11137, Coronavirus OC43 taxID=31631, SARS-CoV-2 taxID=2697049, Paramyxoviridae taxID=11158, Human metapneumovirus taxID=162145, Parainfluenza virus taxID=2905673, Respiratory syncytial virus taxID=12814, Picornaviridae taxID=12058, Rhinovirus taxID=31708, Enterovirus taxID=12059, Orthomyxoviridae taxID=11308, Influenza A taxID=382835, and Influenza B taxID=11520.

### Statistical Analysis

Demographics, symptoms, underlying conditions, and outcomes were described using frequency distributions for categorical variables and medians and interquartile ranges for continuous variables. Subgroup differences were evaluated to compare individuals who were vaccinated with individuals who were not vaccinated, using chi-square tests and Fisher’s exact tests for categorical variables and Wilcoxon rank-sum test for continuous variables.

Prior to testing, the association between clinical factors and mean C_T_ was compared between qRT-PCR testing platforms using ANOVA. There were significant differences between the platforms indicating that platform is an important covariate to control for while modeling factors associated with C_T_.

Missing values were imputed using the chained equations algorithm in the MICE R package^39^ (R Version 4.1.3) to create 10 imputed data sets. Predictive mean matching, logistic regression imputation, and polytomous regression imputation were used for numerical, binary, and multicategory variables, respectively. Results from the 10 linear regression models ran on the imputed datasets were then pooled according to Rubin’s rule to provide an overall estimate for the variables.

Multinomial logistic regressions were used to test the association between demographic characteristics, underlying conditions, vaccine status, and SARS-CoV-2 variant with disease severity (1=Mild [Reference Category], 2=Moderate, 3=Severe, 4=Death). First, unadjusted models tested each variable’s association with disease severity separately. Next, multivariable models were constructed in a step-wise fashion, adding variables in blocks of demographic variables, including age (continuous), sex (0=Female[Reference], 1=Male), and race(0=Black[Reference], 1=White), followed by a block of underlying condition variables, all of which were binary (0=No, 1=Yes; pregnant, chronic lung disease, hypertension, overweight, cardiovascular disease, diabetes, renal disease, liver disease, autoimmune disease, immunocompromised, systemic immunosuppressive therapy/medications), and the final model added a block of SARS-CoV-2-related characteristics including vaccination status (1=Unvaccinated [Reference], 2=Vaccinated, 3=Vaccinated and Boosted), Days Since Most Recent Vaccination/Booster (1=Unvaccinated [Reference], 2=Within past 90 Days, 3=91 – 180 Days Ago, 4=181 – 270 Days Ago, 5=More than 270 Days Ago) and variant (1=Delta[Reference], 2=Omicron). The multivariable models were tested for multicollinearity using variance inflation factor (VIF), though collinearity was not present, so all variables were retained.

All analyses were conducted in R Version 4.1.3 (R Foundation for Statistical Computing, Vienna, Austria).

## ACKNOWLEDGEMENTS

The study was supported by Centers for Disease Control and Preventions contract 75D30121C10084 under BAA ERR 20-15-2997 (to A.P) and by the Emory WHSC COVID-19 Urgent Research Engagement (CURE) Center, made possible by generous philanthropic support from the O. Wayne Rollins Foundation and the William Randolph Hearst Foundation (to A.P.). L.R.C. was supported by Award Number T32AI074492 from the National Institute of Allergy and Infectious Diseases. EIP Surveillance of COVID-19 was funded through the Centers for Disease Control and Preventions Emerging Infections Program [U50CK000485]. The study was supported by the National Institute of Biomedical Imaging and Bioengineering at the National Institutes of Health under award U54 EB027690 (to W.A.L.) and the National Center for Advancing Translational Sciences of the National Institutes of Health under Award Number UL1TR002378 (to W.A.L.). Additional support was provided by the Georgia Clinical & Translational Science Alliance of the National Institutes of Health under Award Number UL1TR002378. The study was supported in part by the Emory Integrated Genomics Core (EIGC) and Emory Integrated Computational Core (EICC), which are subsidized by the Emory University School of Medicine and are one of the Emory Integrated Core Facilities. The study was supported by the Data Analytics and Pediatric Biostatistics Core, Department of Pediatrics, Emory University School of Medicine. The content is solely the responsibility of the authors and does not necessarily represent the official views of the National Institute of Allergy and Infectious Diseases or the National Institutes of Health. The funders played no role in study design, data collection and analysis, decision to publish, or preparation of the manuscript.

## Data Availability

All sequence data are available in NCBI under BioProject PRJNA634356. The GISAID accession number for each sequence is listed in the Supplementary Data File.

## Author contributions

Conceptualization: LRC, ADR, AB, AP

Methodology: LRC, KPO, JJW, AP, RZ, ZZ, RA, LJ, AW

Investigation: LRC, ADR, JJW, DHK, ECF, AP, PN, AT

Validation: ADR, DHK, AP

Visualization: LRC, EW, YL, RML

Data Curation: LRC, ADR, DHK, AB, AA, JHC

Funding acquisition: KPO, WAL, AP

Project administration: LRC, DHK, AP

Supervision: LRC, DAG, AP, RZ

Writing – original draft: LRC, RML, ZZ

Writing – review & editing: DHK, ECF, AB, AP, RML, RZ, RA, JJW, LJ, AW, WAL

Resources: DHK, ECF, DAG, AP, CK, DH

Statistical analyses: RZ, AA, ZZ, LJ, AW

Unrestricted access to all data: KPO, AP

First draft of the manuscript, reviewed it and edited it: LRC, AP

All authors agreed to submit the manuscript, read and approved the final draft and take full responsibility of its content, including the accuracy of the data and the fidelity of the trial to the registered protocol and its statistical analysis.

## SUPPLEMENTARY TABLES

**Table S1.**
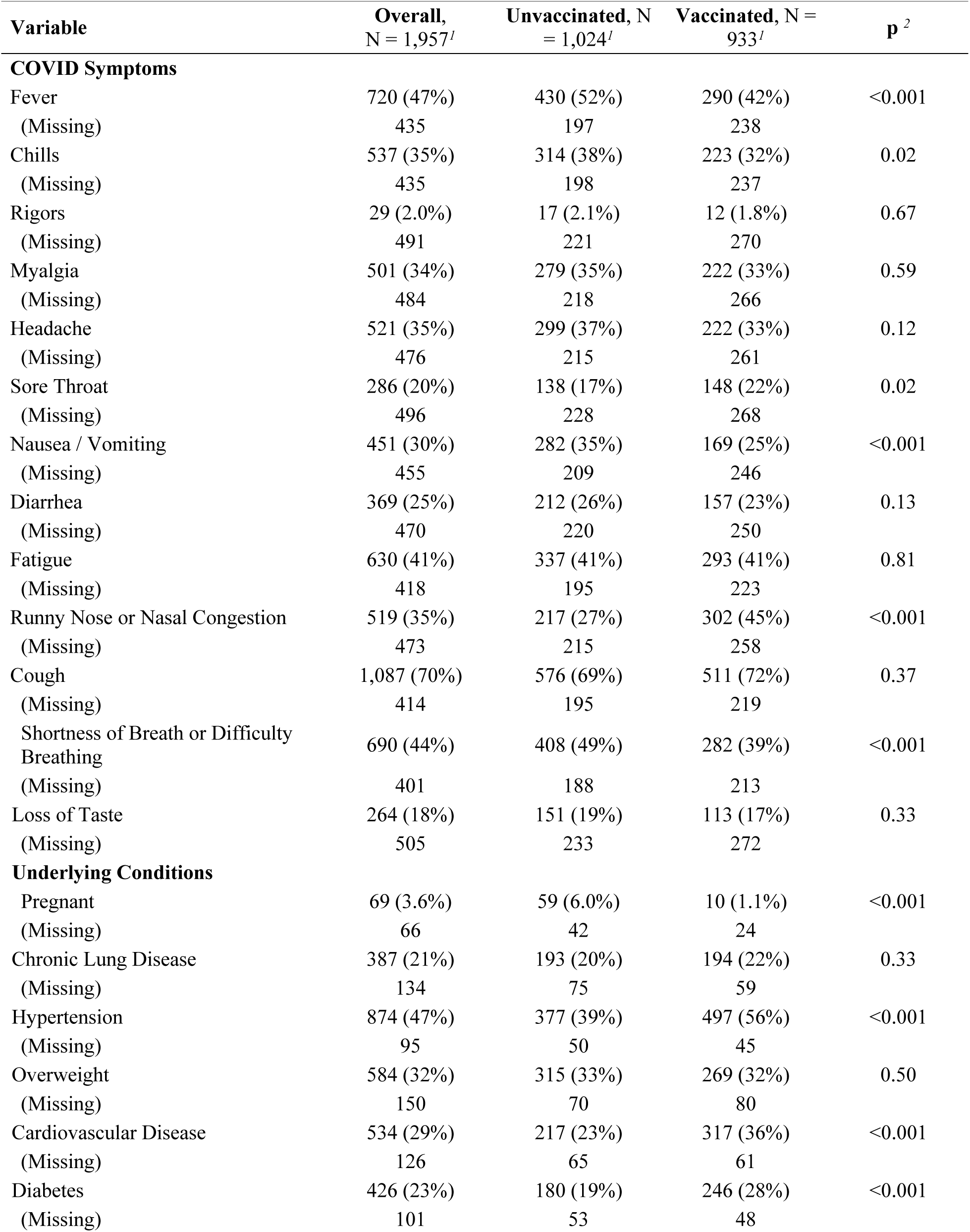
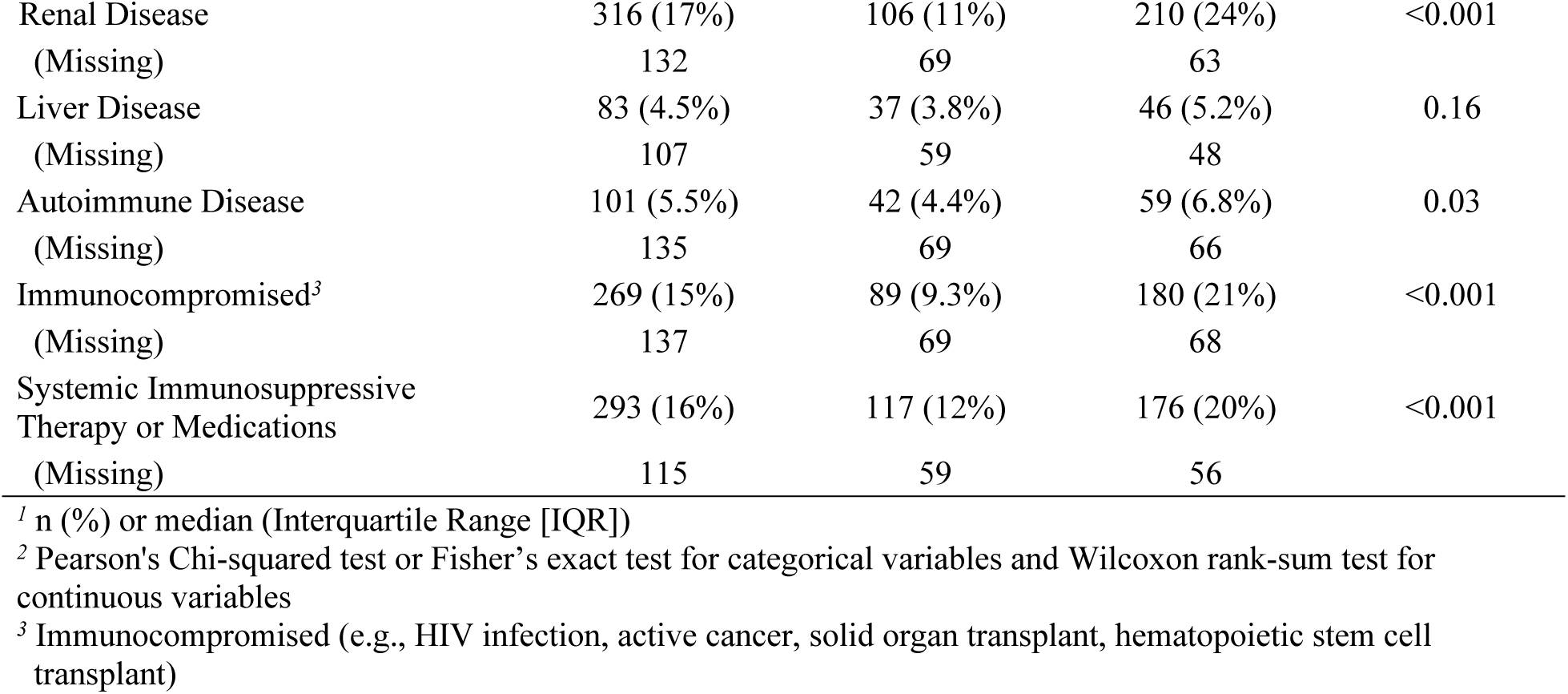
Extended COVID Symptoms and Underlying Conditions by Vaccination Status.

**Table S2.** Circulation of VOC/VOI in Georgia, USA as measured by available sequences on GISAID.

| VOC/VOI | Period variant circulated in GA<sup>1</sup> | Date range of sequences obtained from GISAID | Sequences available on GISAID | Total sequences available from May 2021 to May 2022 (%) |
| --- | --- | --- | --- | --- |
| **Alpha** | 12/10/2020-08/26/2021 | 05/01/2021-08/26/2021 | 1,361 | 2.66 |
| **Beta** | 02/01/2021-05/25/2021 | 05/01/2021-05/25/2021 | 16 | 0.03 |
| **Delta** | 4/29/2021-03/10/2022 | 05/01/2021-03/10/2022 | 28,320 | 55.5 |
| **Gamma** | 03/05/2021-09/24/2021 | 05/01/2021-09/24/2021 | 185 | 0.36 |
| **Lambda** | 02/07/2021-08/05/2021 | 05/01/2021-08/05/2021 | 18 | 0.04 |
| **Mu** | 04/23/2021-09/14/2021 | 05/01/2021-09/14/2021 | 142 | 0.28 |
| **Omicron** | 11/30/2021-4/18/2023 | 11/30/2021-04/18/2023 | 20,759 | 40.6 |
| **Total VOC/VOI** |  |  | **50,801** | **99.46** |
| **Total sequences available on GISAID from May 2021 to May 2022 from the state of Georgia** |  |  | **51,075** | **---** |

<sup>1</sup>Period of circulation based on sequences available on GISAID including Emory Healthcare samples.

**Table S3.** Unadjusted Multinomial Logistic Regression Models Testing the Association between Demographics, Underlying Health Conditions, and COVID-Related Characteristics with Disease Severity.

| Variable | Moderate<sup>1</sup> |  |  | Severe<sup>1</sup> |  |  | Death<sup>1</sup> |  |  |
| --- | --- | --- | --- | --- | --- | --- | --- | --- | --- |
|  | OR<sup>2</sup> | 95% CI<sup>2</sup> | p | OR<sup>2</sup> | 95% CI<sup>2</sup> | p | OR<sup>2</sup> | 95% CI<sup>2</sup> | p |
| **Unadjusted Models** |  |  |  |  |  |  |  |  |  |
| Age | **1.04** | **1.03, 1.05** | **<0.001** | **1.05** | **1.04, 1.05** | **<0.001** | **1.07** | **1.05, 1.09** | **<0.001** |
| Male<sup>3</sup> | **1.40** | **1.13, 1.74** | **<0.01** | 1.30 | 0.95, 1.78 | 0.10 | 1.26 | 0.72, 2.22 | 0.40 |
| White<sup>4</sup> | 0.86 | 0.67, 1.10 | 0.20 | 1.27 | 0.91, 1.77 | 0.20 | 1.04 | 0.57, 1.91 | 0.90 |
| Chronic Lung Disease | **2.07** | **1.59, 2.68** | **<0.001** | **1.95** | **1.35, 2.81** | **<0.001** | **2.65** | **1.44, 4.86** | **<0.01** |
| Hypertension | **2.94** | **2.35, 3.68** | **<0.001** | **4.19** | **2.98, 5.90** | **<0.001** | **11.4** | **5.09, 25.6** | **<0.001** |
| Overweight | **1.45** | **1.15, 1.83** | **<0.01** | **1.58** | **1.14, 2.19** | **0.01** | **2.02** | **1.13, 3.61** | **0.02** |
| Cardiovascular Disease | **2.89** | **2.28, 3.67** | **<0.001** | **3.46** | **2.48, 4.81** | **<0.001** | **4.80** | **2.70, 8.54** | **<0.001** |
| Diabetes | **2.04** | **1.58, 2.63** | **<0.001** | **3.21** | **2.29, 4.50** | **<0.001** | **4.35** | **2.43, 7.80** | **<0.001** |
| Renal Disease | **3.88** | **2.90, 5.19** | **<0.001** | **3.86** | **2.62, 5.67** | **<0.001** | **9.86** | **5.46, 17.8** | **<0.001** |
| Liver Disease | 1.47 | 0.87, 2.50 | 0.20 | **2.36** | **1.25, 4.45** | **<0.01** | **3.08** | **1.16, 8.21** | **0.02** |
| Autoimmune Disease | **1.60** | **1.01, 2.56** | **0.046** | **1.58** | **0.82, 3.03** | **0.20** | **2.45** | **0.93, 6.47** | **0.07** |
| Immunocompromised | **2.65** | **1.97, 3.58** | **<0.001** | **2.44** | **1.61, 3.69** | **<0.001** | **3.69** | **1.93, 7.09** | **<0.001** |
| Systemic Immunosuppressive Therapy/Meds. | **3.49** | **2.61, 4.67** | **<0.001** | **3.24** | **2.18, 4.81** | **<0.001** | **3.48** | **1.79, 6.78** | **<0.001** |
| <u>Vaccination Status</u> |  |  |  |  |  |  |  |  |  |
| Unvaccinated | Ref | — |  | Ref | — |  | Ref | — |  |
| Vaccinated<sup>5</sup> | **0.80** | **0.64, 1.01** | **0.07** | 0.86 | 0.61, 1.21 | 0.40 | 1.00 | 0.54, 1.84 | >0.90 |
| Vaccinated & boosted<sup>5</sup> | **2.28** | **1.52, 3.43** | **<0.001** | **3.24** | **1.95, 5.40** | **<0.001** | **3.69** | **1.58, 8.59** | **<0.01** |
| <u>Days Since Most Recent Vaccination/Booster</u> |  |  |  |  |  |  |  |  |  |
| Unvaccinated | — | — |  | — | — |  | — | — |  |
| Within past 90 Days<sup>5</sup> | 1.34 | 0.86, 2.08 | 0.20 | 1.68 | 0.93, 3.03 | 0.09 | 2.81 | 1.17, 6.79 | 0.02 |
| 91 - 180 Days Ago<sup>5</sup> | 0.98 | 0.74, 1.29 | 0.90 | 0.92 | 0.60, 1.40 | 0.70 | 1.14 | 0.55, 2.37 | 0.70 |
| 181 - 270 Days Ago<sup>5</sup> | 0.72 | 0.51, 1.01 | 0.06 | 0.74 | 0.45, 1.24 | 0.30 | 0.71 | 0.27, 1.89 | 0.50 |
| More than 270 Days Ago<sup>5</sup> | 1.18 | 0.72, 1.94 | 0.50 | 2.49 | 1.43, 4.36 | <0.01 | 1.91 | 0.64, 5.72 | 0.20 |
| <u>Lineage</u> |  |  |  |  |  |  |  |  |  |
| Delta | Ref | — |  | Ref | — |  | Ref | — |  |
| Omicron<sup>6</sup> | **1.51** | **1.09, 2.07** | **0.01** | **2.25** | **1.46, 3.48** | **<0.001** | **4.55** | **2.25, 9.20** | **<0.001** |

<sup>1</sup> "Mild" is the Reference Category
<sup>2</sup> OR = Odds Ratio, CI = Confidence Interval, Ref = Reference Level
<sup>3</sup> Compared to "Female"
<sup>4</sup> Compared to "Black"
<sup>5</sup> Compared to "Unvaccinated individuals"
<sup>6</sup> Compared to "Delta"

## SUPPLEMENTARY FIGURES

**Figure S1.**
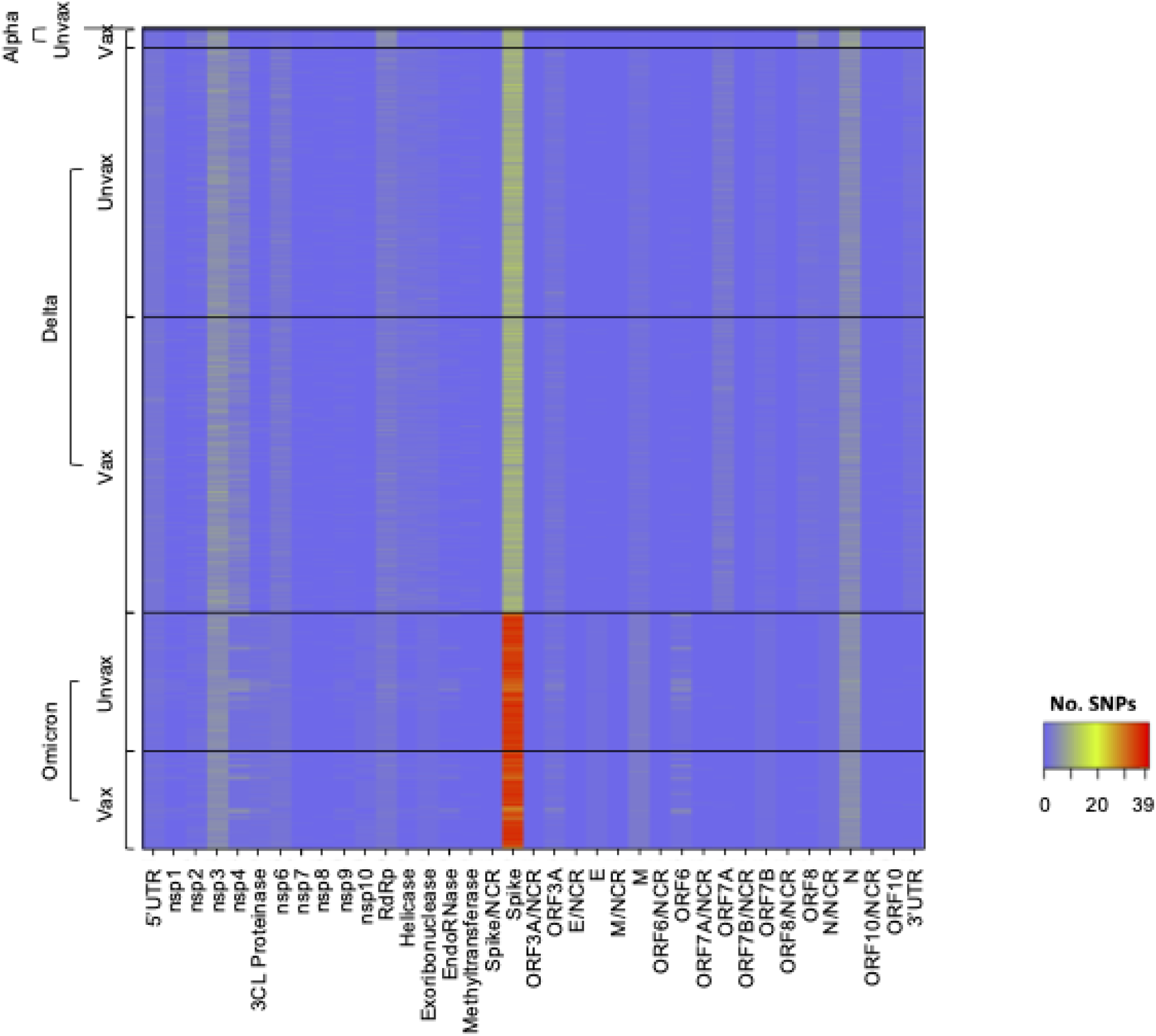
Mutations across SARS-CoV-2 genes. Alpha, delta, and omicron sequences were aligned to Wuhan/Hu-1/2019. For each variant, samples from vaccinated (“vax”) and unvaccinated (“unvax”) individuals are labeled. Each square in the heatmap represents the number of single nucleotide polymorphisms (SNPs) in each gene, labeled on the x-axis. Each row represents one sample.

**Figure S2.**
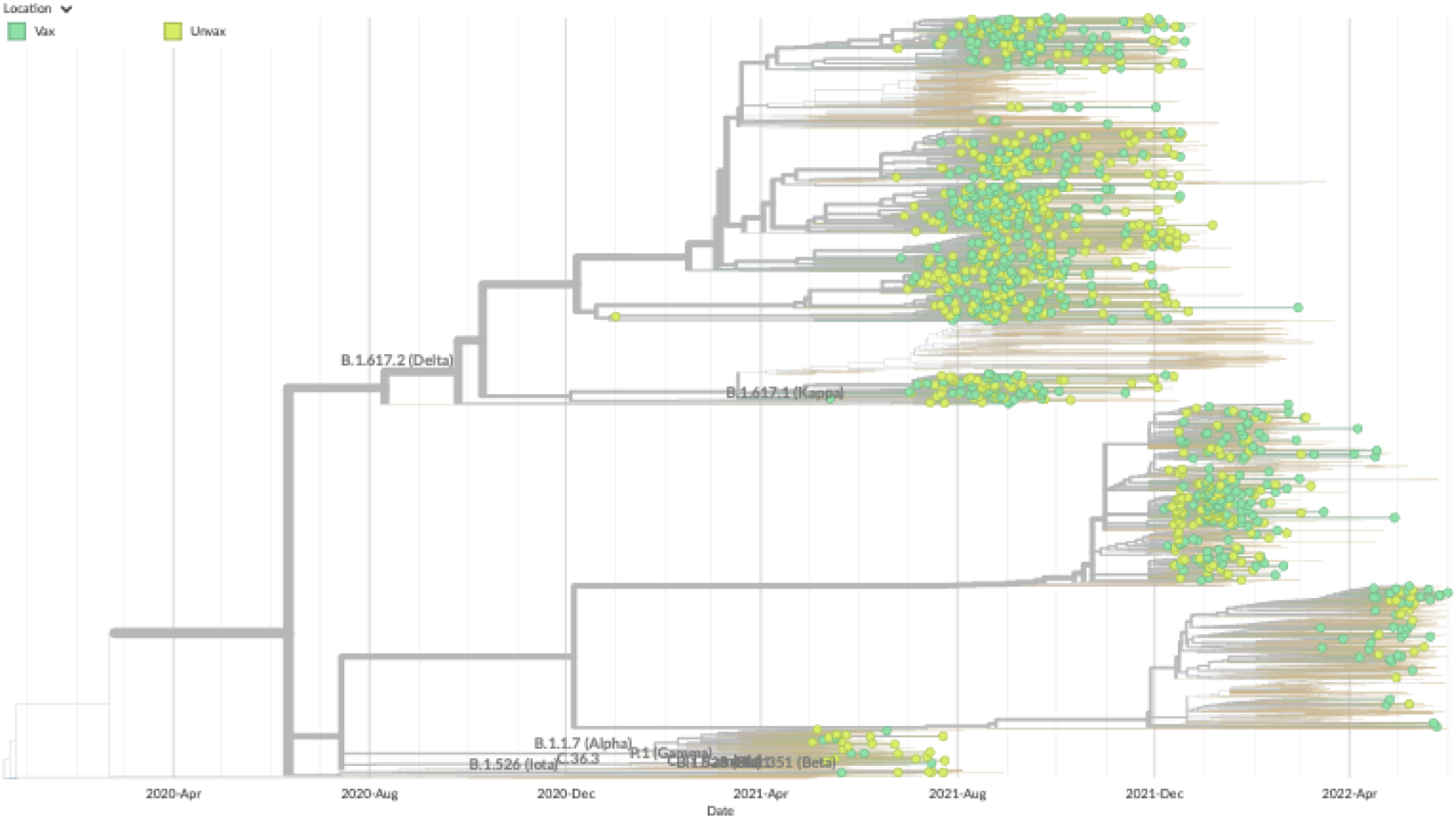
Phylogenetic analysis does not reveal differences in SARS-CoV-2 sequences from vaccinated and unvaccinated individuals. Maximum likelihood tree containing sequences from vaccinated (green) and unvaccinated (yellow) individuals in the context of 2000 global sequences from GISAID (orange) selected by a custom Nextstrain subsampling scheme and rooted to NC_045512.

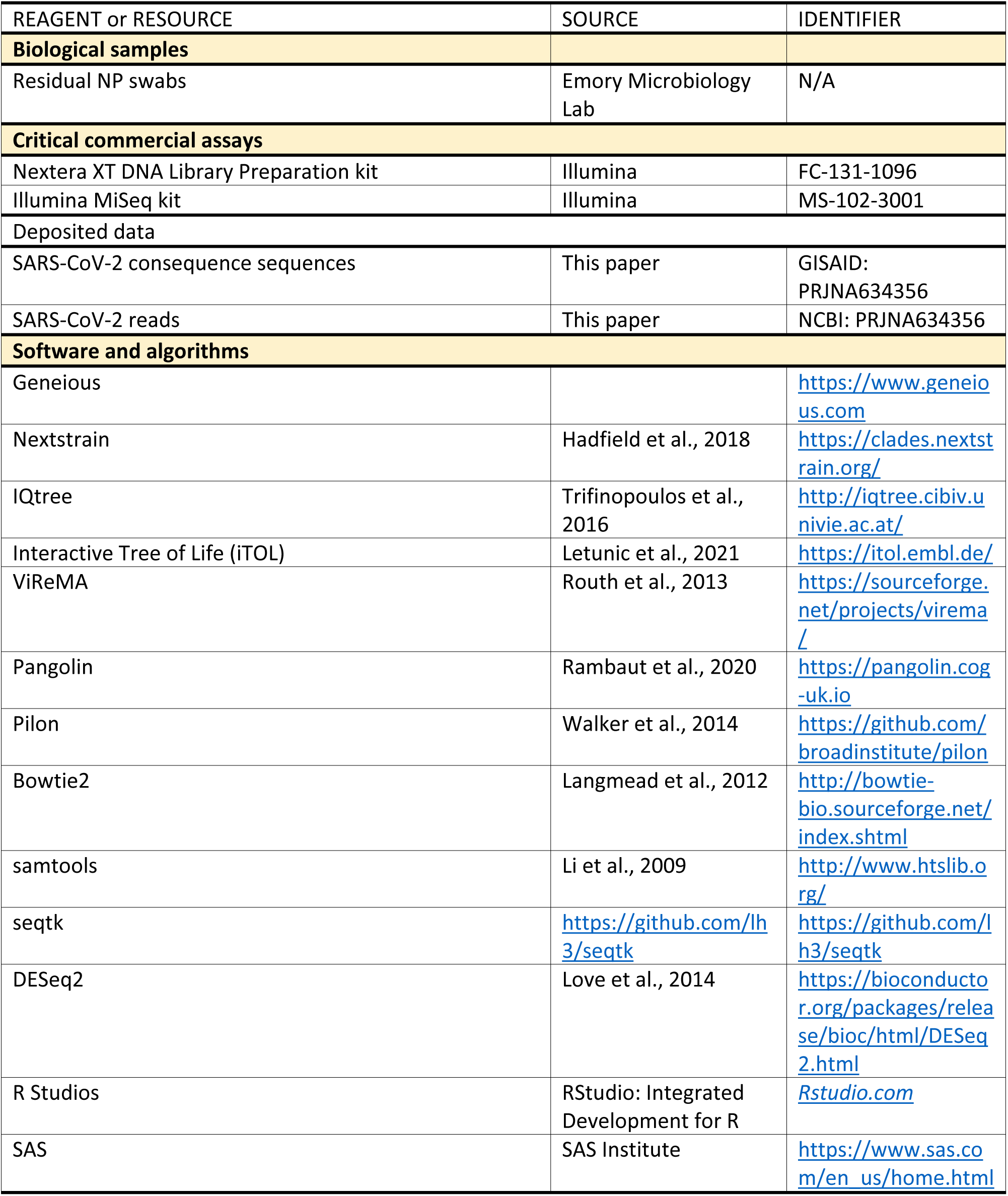
KEY RESOURCES TABLE.

